# Microstructure predicts impulsive and compulsive behaviour following subthalamic stimulation in Parkinson’s disease

**DOI:** 10.64898/2026.04.13.26350763

**Authors:** Philipp A. Loehrer, Laura Witt, Maja Nagel, Lijiao Chen, Alexander Calvano, Miriam H. A. Bopp, Alexandra Rizos, Maxine Hillmeier, Julius Wichmann, Christopher Nimsky, K. Ray Chaudhuri, Haidar S. Dafsari, Lars Timmermann, David J. Pedrosa, Marcus Belke

## Abstract

**Background:** Subthalamic deep brain stimulation (STN-DBS) represents an established therapeutic intervention for advanced Parkinson’s disease (PD), alleviating motor and non-motor symptoms. However, impulse control disorders (ICDs) present a complex challenge, with some patients experiencing postoperative improvements while others develop treatment induced impulsive-compulsive behaviours (ICB). The mechanisms determining these variable outcomes remain poorly understood, highlighting the need to predict postoperative ICB outcomes.

**Methods:** This prospective open-label study aimed to identify microstructural markers associated with postoperative changes in impulsive-compulsive behaviour following STN-DBS. Thirty-five patients underwent diffusion MRI and clinical evaluations preoperatively and six months postoperatively. A whole-brain voxel-wise analysis utilising diffusion tensor imaging (DTI) and neurite orientation dispersion and density imaging (NODDI) was conducted to explore associations between microstructural metrics and changes in the Questionnaire for Impulsive-Compulsive Disorders in Parkinson’s Disease-Rating Scale (QUIP-RS).

**Results:** Intact microstructure in frontolimbic WM tracts, including the cingulum, insular cortex connections, and major association fibres, was associated with greater postoperative reductions in impulsive-compulsive symptoms. Conversely, intact microstructure in specific grey matter areas including paracingulate gyrus, insular cortex, and precentral gyrus were associated with lower reductions or increases in postoperative ICB.

**Conclusion:** These findings demonstrate that preoperative microstructural integrity within frontolimbic circuits and executive control networks associates with susceptibility to treatment-emergent impulsive-compulsive behaviours following STN-DBS. The convergent evidence from multiple diffusion metrics suggests that diffusion MRI may serve as a valuable tool for identifying patients at risk for developing ICB, potentially enhancing preoperative counselling and enabling targeted behavioural monitoring strategies.

## Introduction

Parkinson’s disease (PD) is characterised by both motor and non-motor symptoms which significantly diminish patients’ quality of life.^1^ Subthalamic deep brain stimulation (STN-DBS) is a well-established treatment for motor deficits and has also been shown to alleviate many non-motor manifestations, including mood, sleep, and anxiety.^2–5^ Among these non-motor symptoms, impulse control disorders (ICDs) represent a particularly challenging and heterogeneous domain which is highlighted as a key “vital” in the management of Parkinson’s in the newly released “The Parkinson’s Companion app” to ensure its management in routine practice.^6^ In fact, the effects of STN-DBS on ICDs are complex and variable: some patients with preexisting ICDs experience postoperative improvements – partially attributed to the reduction of dopaminergic medication – while others without prior ICDs develop new or worsened impulsive-compulsive behaviours (ICB), possibly due to the spread of stimulation within cortico-subcortical networks.^7^ ^8^ These divergent outcomes suggest that the stimulated regions, their microstructural integrity, and the integrity of the connected pathways play a pivotal role in non-motor responses to STN-DBS.^9^ ^10^ Indeed, recent diffusion MRI studies have demonstrated that preserved microstructural integrity within the basal ganglia-thalamo-cortical circuit correlates with favourable non-motor outcomes.^9^ Advanced modelling techniques such as neurite orientation dispersion and density imaging (NODDI) offer greater sensitivity than conventional diffusion tensor imaging (DTI) by estimating voxel-wise neurite density index (NDI) – quantifying the density of axons and dendrites – and orientation dispersion index (ODI) – assessing the dendritic arborisation.^11–14^ In the present study, we leverage whole-brain NODDI- and DTI-derived metrics to determine whether preoperative microstructural features correlate with postoperative changes in impulsivity, as measured by the Questionnaire for Impulsive-Compulsive Disorders in Parkinson’s Disease-Rating Scale (QUIP-RS).

## Methods

### Participants

This ongoing observational study enrolled 37 patients with PD upon written informed consent. Inclusion criteria comprised indication for DBS surgery due to advanced PD following international guidelines.^15^ Study exclusion criteria were pathological MRI findings, reduced auditory or visual functions, or simultaneous presence of neurological or psychiatric conditions. Due to missing follow-up data, two participants were excluded resulting in a cohort of 35 patients. As this study is part of a research initiative, the cohort partially overlaps with that of previous publications.^9^ ^14^ However, the present analysis addresses a specific non-motor outcome. The study was approved by the ethics committee of the Philipps-University of Marburg (study-number:155/17) and adhered to the principles of the Declaration of Helsinki.

### Clinical Assessments

Patients underwent evaluations preoperatively and six months postoperatively. While baseline assessments were conducted in the ON medication state, follow-up evaluations were conducted in the ON-medication/ON-stimulation state. Here, standardised case report forms were used to gather demographic and the following clinical data: the QUIP-RS, the Parkinson’s Disease Questionnaire 8 (PDQ-8), and the Levodopa Equivalent Daily Dose (LEDD). The QUIP-RS features a 4×7 format consisting of four main questions that focus on commonly reported thoughts, urges/desires, and behaviours related to ICB.^16^ These questions are applied across seven items: the first four items cover ICD (such as gambling, buying, sexual, and eating behaviours), items five and six evaluate other compulsive behaviours (punding and hobbyism), and item seven addresses compulsive medication use. Each of the four primary questions includes all seven items, which are rated on a five-point Likert scale ranging from 0 (never) to 4 (very often). Consequently, the total QUIP-RS score can range from 0 to 112. Clinically relevant ICDs were defined as QUIP-RS subscale scores ≥ 10 (items 1-4) and ≥ 7 (items 5-6).^16^ The PDQ-8 assesses quality of life across eight dimensions in PD and provides a summary index (SI) ranging from 0 (no impairment) to 100 (maximum impairment). LEDD calculations followed international standards described by Jost et al.^17^ Postoperative medication titration and stimulation adjustments adhered to standardised protocols to ensure consistency across patients.^18^

### MRI Data Acquisition and Processing

Patients with PD were scanned on a 3T Trio scanner (Siemens, Erlangen, Germany) in the ON-medication state at preoperative baseline. All MRI scans were acquired at the University of Marburǵs Core Unit Brain Imaging. The imaging protocol is detailed in the supplementary material. All scans were reviewed for motion, ghosting, and high-frequency and/or wrap around artifacts. To rule out postoperative complications as intracranial bleeding, edema, or lead displacements, postoperative computed tomography (CT) was performed. There were no postoperative complications observed on imaging.

### Image Processing

The structural MRI data analysis was performed using the FreeSurfer software version 7.1.1. (http://surfer.nmr.mgh.harvard.edu).^19^ Skull stripping, automated Talairach transformation, cortical and subcortical segmentation, intensity normalisation, grey/WM boundary tessellation, topology correction, and surface deformation based on intensity gradients were steps of the processing pipeline. Analysis of the diffusion-weighted imaging data was performed by using the FMRIB Software Library (FSL) analysis suite version 6.0.5.2. (https://fsl.fmrib.ox.ac.uk/fsl).^20^ To correct the raw diffusion volumes for eddy-current distortions and artifacts the volumes were registered and resampled to the first b0 volume. Linear regression was used to calculate diffusion tensors for each voxel to derive Fractional Anisotropy (FA) maps. Furthermore, NODDI-DTI (a modification of NODDI) was used to extract NDI and ODI. Visual inspection of the b0 images was performed to ensure that there are no alterations in the images that could affect the results. Using a boundary-based registration method, the first b0 image of each scan was registered to the T1-weighted structural images, yielding an affine matrix. T1-derived segmentations and brain masks were transformed into diffusion space using the inverse of the affine matrix. To transform masked FA-, NDI- and ODI-maps into the MNI152 standard space, linear and non-linear transformations were used. Voxels of brain tissue not present throughout all subjects were excluded from further analysis.

The Lead-DBS toolbox with default parameters (https://www.lead-dbs.org/) was used to verify lead placement.

### Statistical Analysis

Statistical analyses of clinical outcomes were performed using MATLAB R2020b (The MathWorks, Inc.). Changes from baseline to six-month follow-up (6-MFU) were analysed using T-tests or Wilcoxon signed-rank tests, depending on the normality assessments conducted via Shapiro-Wilk tests. The Benjamini-Hochberg method was applied to control the false discovery rate (FDR), and statistical significance was determined based on FDR-adjusted p-values at a threshold of p_adj_<.05. Effect sizes were calculated in accordance with Coheńs convention. Statistical voxelwise analysis of imaging data was performed using a generalised linear model including the covariates age and disease duration as described previously.^9^ Here, percentage differences between baseline and follow-up values for the QUIP-RS Total score were calculated. A permutation-based approach, based on the Analysis of Functional Neuro Images (AFNI) null-z simulator, was used to correct for multiple comparisons employing 12,000 simulations under the null hypothesis. Clusters were formed using a threshold of p<.01 and a clusterwise p-value was calculated. Results were accepted as significant with a clusterwise p<.05.

## Results

### Clinical outcomes

At the 6-MFU, the following outcomes improved: PDQ-8 SI (p<.001, Cohen’s d=.71), LEDD (p<.001, Cohen’s d=1.22), and LEDD of dopamine agonists (p<.001, Cohen’s d=.64). No significant change in QUIP-RS Total was observed (p=.94, Cohen’s d=-.07). In a subgroup of patients with clinically relevant preoperative ICD, QUIP-RS Total (p=.003, Cohen’s d=.71) and the subscores Hobbyism (p=.018, Cohen’s d=.8), and Punding (p<.001, Cohen’s d=.87) improved. Longitudinal changes of clinical outcomes are reported in **Table 1** and **Figure 1**.

**Table 1:** Baseline characteristics and outcomes at baseline and 6-month follow-up.

|  | *n* | *M* | *SD* |
| --- | --- | --- | --- |
| Age [y] | 35 | 58.7 | 7.4 |
| Disease duration [y] | 35 | 8.9 | 4.5 |
| Sex (female/male) [%] | 35 | 9/26 | [22.9/77.1] |

**Table 1:** Demographic characteristics and outcome parameters at baseline and 6-months follow-up. Reported p-values are corrected for multiple comparisons using Benjamini-Hochberg's method. Bold font highlights significant results, $p < .05$ .
|  | Baseline |  |  | 6-MFU |  |  | Baseline vs. 6-MFU |  |
| --- | --- | --- | --- | --- | --- | --- | --- | --- |
|  | *n* | *M* | *SD* | *n* | *M* | *SD* | *p* | *Cohen's d* |
| QUIP-RS Total | 35 | 17.9 |  | 35 | 19 |  | .94 | -.07 |
| Gamble | 35 | .46 | 1.2 | 35 | .74 | 1.4 | .20 | -.22 |
| Sex | 35 | 2.7 | 3.3 | 35 | 2.7 | 2.8 | .95 | -.01 |
| Buy | 35 | 2.7 | 3.1 | 35 | 2.6 | 2.5 | .50 | .05 |
| Eat | 35 | 3.4 | 3 | 35 | 4.3 | 3.4 | .16 | -.29 |
| Hobby | 35 | 3.5 | 3.4 | 35 | 3.3 | 2.7 | .57 | .07 |
| Punding | 35 | 2.5 | 2.9 | 35 | 2.4 | 2.9 | .40 | .06 |
| DDS | 35 | 2.7 | 3.7 | 35 | 2.9 | 4.2 | .91 | -.07 |
| QUIP-RS Total clinically relevant | 15 | 32.5 | 10.7 | 15 | 23.7 | 14 | **.003** | **.71** |
| Gamble | 15 | .73 | 1.6 | 15 | .73 | 1.4 | 1 | 0 |
| Sex | 15 | 4.5 | 3.7 | 15 | 3.1 | 2.8 | .09 | .44 |
| Buy | 15 | 5 | 3.3 | 15 | 4 | 2.4 | .17 | .35 |
| Eat | 15 | 5.9 | 2.6 | 15 | 5.9 | 3.4 | .95 | .02 |
| Hobby | 15 | 6.4 | 2.9 | 15 | 4.3 | 2.1 | **.018** | **.8** |
| Punding | 15 | 5.1 | 2.7 | 15 | 2.7 | 2.8 | **<.001** | **.87** |
| DDS | 15 | 4.8 | 4.4 | 15 | 3 | 4.2 | .07 | .42 |
| PDQ-8 SI | 35 | 32.9 | 15.1 | 35 | 22.2 | 14.6 | **<.001** | **.71** |
| LEDD [mg] | 35 | 965.4 | 403.8 | 35 | 543.5 | 287.9 | **<.001** | **1.22** |
| LEDD-DA [mg] | 35 | 261.3 | 139.1 | 35 | 176.9 | 125.3 | **<.001** | **.64** |

**Figure 1.**
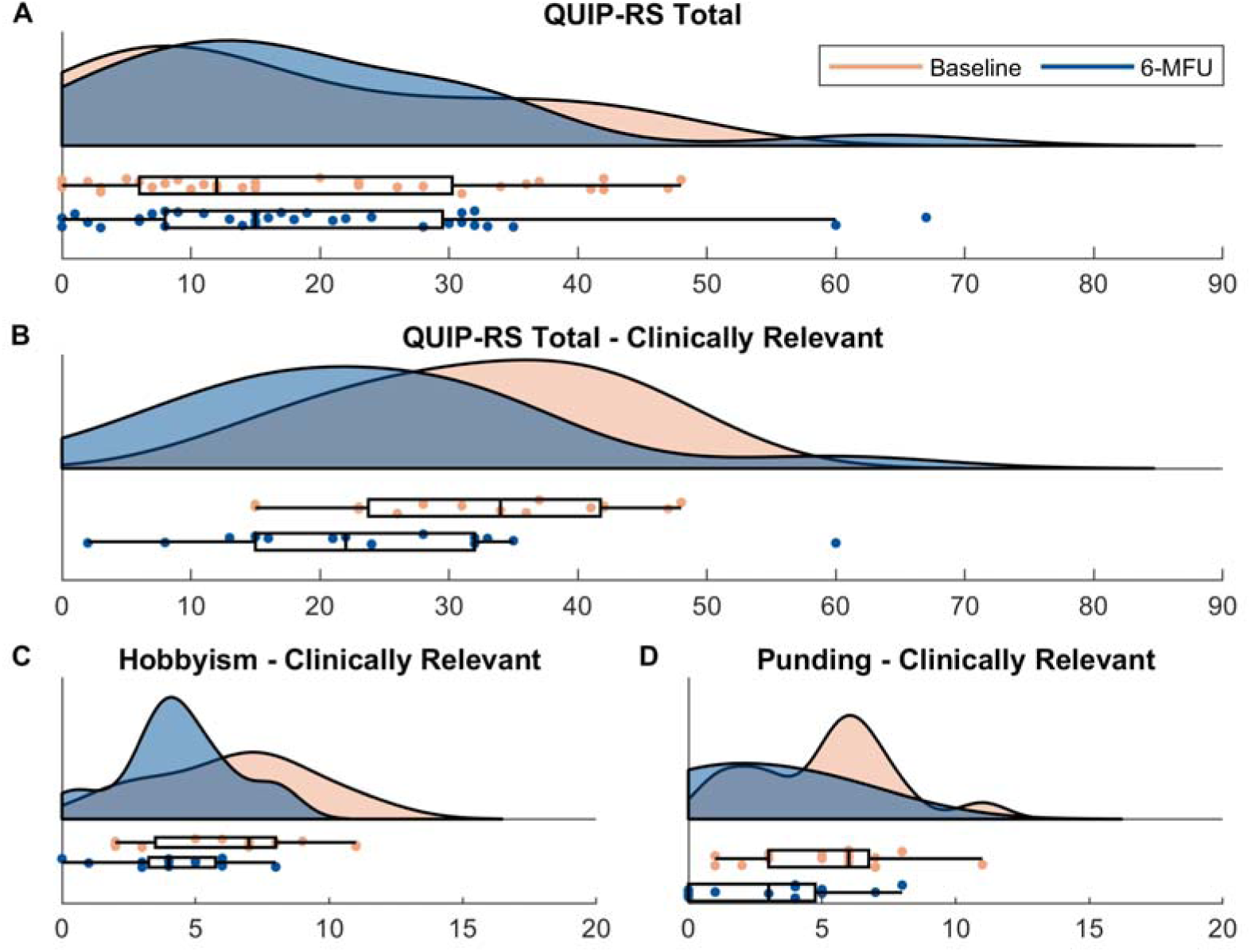
Visualisation of baseline and 6-month follow-up values of QUIP-RS for the whole sample **(A)**. Panels B-D display baseline and 6-MFU values for QUIP-RS for patients with clinically relevant ICD (**B**) and the subscores hobbyism (**C**) and punding (**D**). Center line indicates the median, box limits represent upper and lower quartiles and whiskers indicate most extreme data points not considered outliers.

### Fractional Anisotropy and Postoperative Impulsive-Compulsive Outcomes in PD

Whole □ brain voxel-wise analyses revealed significant FA clusters associated with postoperative change in QUIP-RS (cf. **Supplementary Table 1**, **Supplementary Figure 1**). In a left insular cluster extending into the uncinate fasciculus and inferior fronto-occipital fasciculus, higher preoperative FA was associated with a lower reduction in postoperative QUIP-RS (N4, p<.001). A second cluster in the right cerebellar Crus I/Lobule VI showed that greater FA was also associated with a lower reduction in postoperative QUIP-RS (N7, p<.001). In contrast, a positive FA cluster within the left cingulum revealed that higher preoperative FA was associated with a greater reduction in postoperative QUIP-RS (P1, p<.001).

### Orientation Dispersion Index and Postoperative Impulsive-Compulsive Outcomes in PD

Whole□brain voxel-wise analyses identified multiple significant ODI clusters, all showing negative associations with ΔQUIP□RS (cf. **Supplementary Table 2**, **Figure 2**). The strongest effect was seen in a cluster spanning the cingulum (N7, p<.001), indicating that lower preoperative ODI in this tract associates with greater postoperative reductions in QUIP-RS. Similarly, lower ODI in WM (WM) regions adjacent to the insular cortex and its connections with the superior longitudinal and uncinate fasciculi (N9, p<.001), WM regions adjacent to the precuneous and the cingulum (N15, p<.001), as well as the uncinate fasciculus adjacent to the frontal orbital cortex (N11, p<.001) were associated with larger postoperative reductions in QUIP□RS. Three clusters in the frontal pole – all involving the forceps minor and one extending into the superior frontal gyrus (N2, N3, N8, all: p<.001) – and a cluster in the WM adjacent to the superior parietal lobule and the cingulum (N1, p<.001) were further associated with favourable postoperative QUIP□RS outcomes. Additional associations were present adjacent to the postcentral gyrus within the corticospinal tract and superior longitudinal fasciculus (N5, p<.001), the superior longitudinal fasciculus adjacent to the precentral gyrus (N18, p<.001), and within the inferior fronto□occipital fasciculus (N6, p<.001). Across clusters, ODI effects predominantly localised to WM tracts including the cingulum, forceps minor, and superior longitudinal fasciculus.

**Figure 2.**
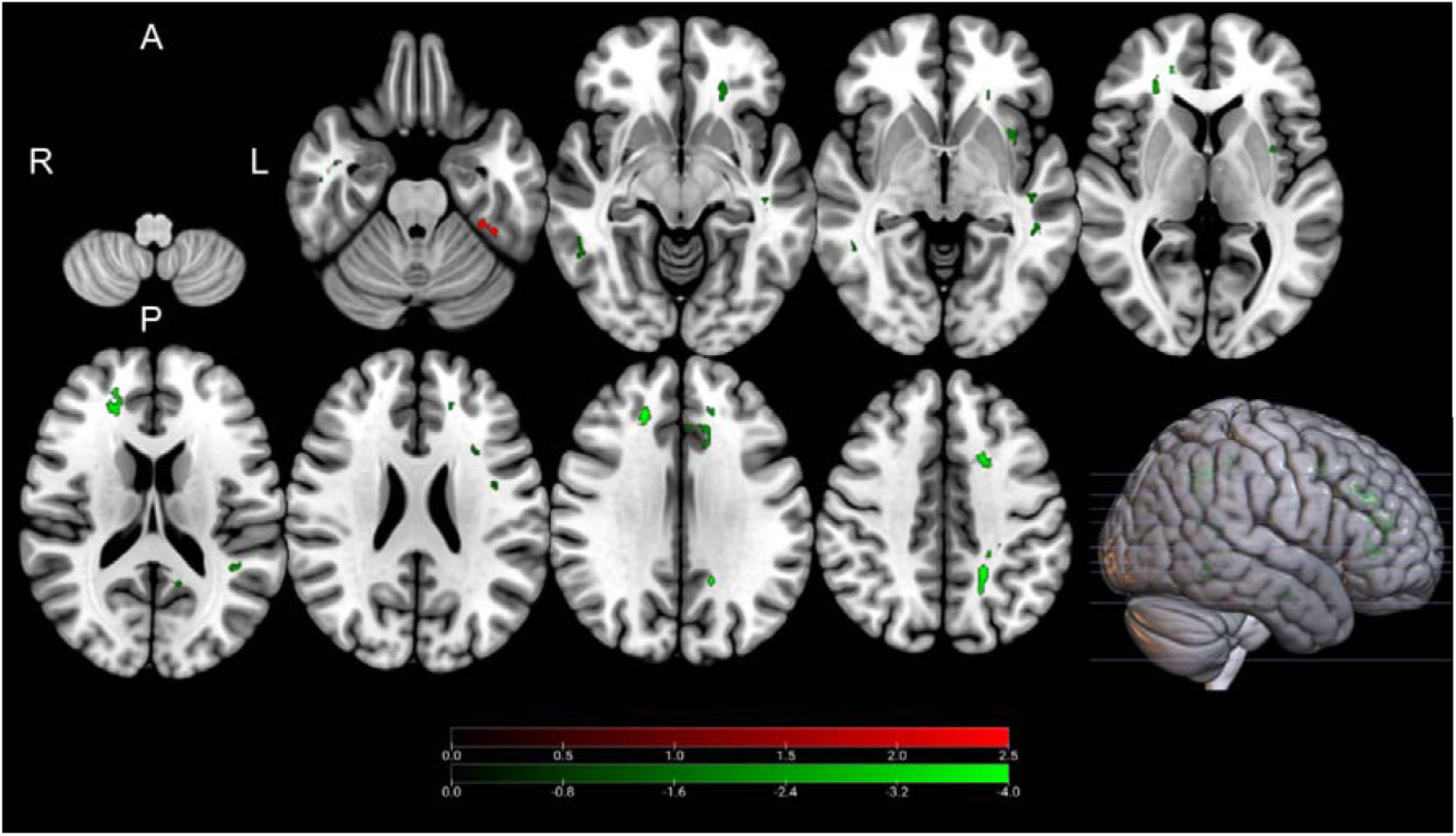
Clusters with a positive (green) and negative (red) association between PD patients’ ODI-values and postoperative change in QUIP-RS, as revealed by the whole brain analysis. Clusterwise p-values were corrected for multiple comparisons using a permutation-based approach and shown as the negative decadic logarithm of the p-value.

### Neurite Density Index and Postoperative Impulsive-Compulsive Outcomes in PD

Preoperative neurite density, as assessed by NODDI□derived NDI, had a strong association with postoperative change in QUIP-RS (cf. **Supplementary Table 3**, **Figure 3**). Whole□brain voxel□wise analysis revealed negative clusters within the grey matter (GM) in which higher NDI was consistently associated with lower postoperative decreases – and even increases – in QUIP□RS. The clusters were located in the bilateral paracingulate gyrus (left: N7, p<.001; right N3, p<.001), the insular cortex extending to the uncinate fasciculus (N4, p<.001), the precentral gyrus (N5, p<.001), right cerebellar lobuli I-IV, right brainstem nuclei within the mesencephalon (N11 p<.001), and the left putamen (N13, p<.001).

**Figure 3.**
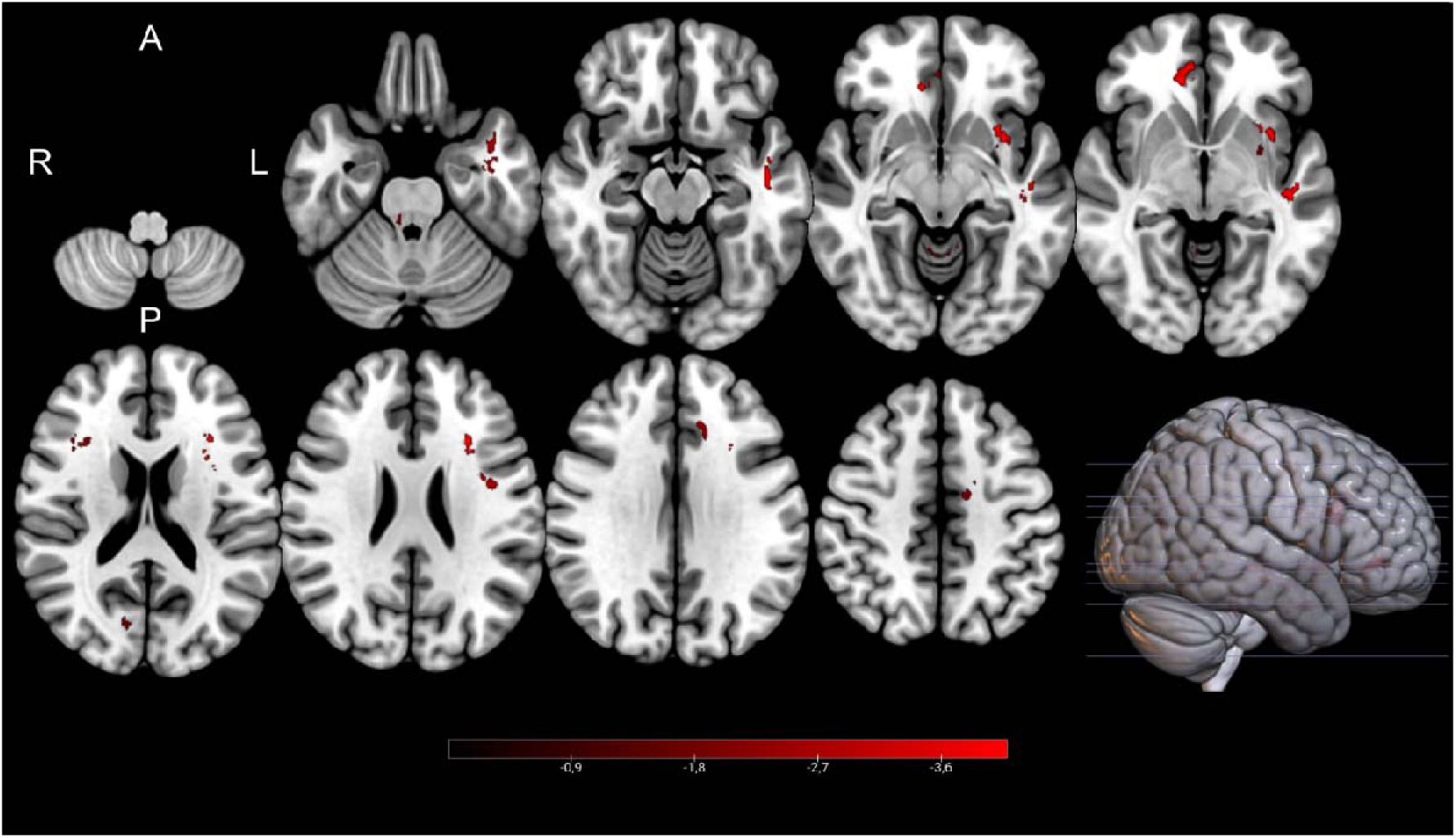
Clusters with a negative (red) association between PD patients’ NDI-values and postoperative change in QUIP-RS, as revealed by the whole brain analysis. Please note that no positive clusters were detected. Clusterwise p-values were corrected for multiple comparisons using a permutation-based approach and shown as the negative decadic logarithm of the p-value.

## Discussion

In the present study, we employed advanced diffusion MRI techniques – including NODDI – to examine how preoperative cerebral microstructure influences susceptibility to treatment-emergent ICBs following STN-DBS in patients with PD. Our results reveal distinct patterns of white and grey matter integrity that are associated with postoperative ICB outcomes, highlighting the potential for diffusion-based markers to inform preoperative risk stratification and individualised patient counselling.

### Microstructure as a predictor for postoperative changes in ICB

STN-DBS is an established therapeutic intervention for advanced PD alleviating motor and non-motor symptoms.^21–24^ Besides the positive effects of STN-DBS, the postoperative emergence of ICB constitutes a clinically significant and pathophysiologically complex phenomenon that requires intensive clinical research.^8^ ^25^ The clinical impact of postoperative ICB is not only a burden for the affected but also for the caregivers.^25^ Behavioural manifestations frequently result in profound disruption of psychosocial functioning, financial instability (pathological gambling, compulsive shopping), hypersexuality, and binge eating.^25^ The cascading effects on family dynamics and caregivers stress represent an underappreciated dimension of disease burden that significantly compromises therapeutic outcomes and long term prognosis.^25^ Currently, a significant deficit in comprehensive knowledge regarding the predictive factors that determine which patients benefit from subthalamic stimulation versus those who exhibit an elevated risk for developing postoperative ICDs exists. This knowledge gap represents a considerable challenge for preoperative patient counselling and therapeutic planning.^8^ ^9^ Therefore, imaging derived markers including metrics of cerebral microstructure have been suggested to predict clinical outcomes and support preoperative patient counselling, stratification, and planning.^9^ ^26^

### Microstructural vulnerability patterns in frontolimbic circuits

The most striking finding of our analysis is the consistent involvement of frontolimbic WM tracts and frontostriatal circuits in predicting QUIP-RS changes. Higher preoperative FA-values in the cingulum were associated with higher reductions in postoperative ICB, indicating that intact WM integrity in these regions confers protection against treatment-emergent ICB. Conversely, higher FA in the left paracingulate gyrus and insular cortex was associated with worse postoperative outcomes, suggesting region-specific patterns of vulnerability that may reflect distinct pathophysiological mechanisms within frontolimbic circuits.

In particular, the WM adjacent to the insula emerged as a critical structure across multiple diffusion metrics, showing significant associations in ODI analyses with beneficial ICB outcomes. This convergent evidence underscores the importance of WM integrity in regions connecting the insula to other brain areas involved in reward evaluation networks and emotional regulation. Previous neuroimaging studies in patients with PD who experience ICBs have consistently implicated altered connectivity involving insular regions in reward processing abnormalities.^27^ Furthermore, WM tracts adjacent to the insula serve as crucial pathways connecting visceral processing areas with conscious awareness networks, making them particularly relevant for understanding the neural basis of impulsive behaviours.^28–30^ The insula serves as a critical neuroanatomical centre that integrates ascending visceral afferent signals with higher-order cognitive and emotional processing networks.^30^ This integration is essential for the generation of conscious awareness regarding internal bodily states, emotional valence and motivational drives.^30^ In the context of impulse control, the insula processes interoceptive information related to reward anticipation, autonomic arousal, and somatic markers that typically inform decision-making processes.^28^ ^31^ Disruption of this integration compromises the ability to consciously recognise and appropriately respond to internal cues that would normally facilitate behavioural regulation.^30^ ^31^

### White matter tract integrity and impulse control

The involvement of major association tracts, particularly the superior longitudinal fasciculus, uncinate fasciculus, and inferior fronto-occipital fasciculus, provides insights into the anatomical substrates underlying ICB susceptibility. The superior longitudinal fasciculus connects frontoparietal regions involved in executive control and attention, while the uncinate fasciculus links orbitofrontal and temporal regions critical for emotional regulation and reward processing.^32–34^ Compromised integrity of these tracts, as reflected by altered NODDI parameters, may disrupt the normal balance between impulsive drives and cognitive control mechanisms.^12^ ^35^

Particularly noteworthy is the consistent involvement of the cingulate WM across multiple analyses. The cingulum, a major limbic tract connecting anterior cingulate, posterior cingulate, and hippocampal regions, forms a critical WM component of circuits involved in emotional processing and behavioural control.^36^ The finding that lower ODI values in paracingulate WM regions predispose to better outcomes indicates that the microstructural integrity of cingulate-associated WM tracts is crucial for maintaining impulse control after STN-DBS.

### Frontoparietal control networks and executive function

The extensive involvement of WM regions adjacent to frontoparietal areas, including regions near the superior parietal lobule, frontal poles, and pre-/postcentral gyri, highlights the importance of WM tracts connecting executive control networks in ICB development.^37–39^ These WM regions are fundamental for maintaining connectivity within the central executive network, responsible for cognitive control, working memory, and decision-making processes.^38^ ^40–42^ The consistent negative associations between NODDI parameters and QUIP-RS outcomes in these WM areas suggest that patients with compromised frontoparietal WM microstructure may have reduced capacity for efficient inter-regional communication, rendering them more susceptible to impulsive behaviours following DBS surgery.^11^ ^43^

The involvement of the forceps minor, connecting bilateral frontal regions, further emphasizes the role of interhemispheric frontal connectivity in impulse control.^44^ Disrupted frontal connectivity may impair the ability to inhibit inappropriate responses and maintain goal-directed behaviour, key components of executive function that are compromised in ICBs.^45^

### Grey matter microstructure and inhibitory control vulnerability

Our findings of elevated NDI in limbic and associative cortical regions among patients with worse postoperative impulsivity outcomes demand further consideration. To date, no studies have directly examined the association between cerebral microstructure and postoperative changes in impulsivity following STN-DBS, necessitating the integration of evidence from related neuropsychiatric conditions to contextualise our findings. Comparable patterns of microstructural abnormality have been documented in conditions characterised by impaired inhibitory control and dysregulated reward processing. In young adult binge drinkers, Morris et al. demonstrated that higher orientation dispersion in the ventral striatum was positively correlated with binge severity, suggesting that increased dendritic complexity in reward-related structures may reflect neuroplastic adaptations associated with compulsive behavioural patterns.^46^ Similarly, NODDI studies in attention-deficit/hyperactivity disorder (ADHD) have revealed associations between higher neurite density in frontostriatal circuits and impulsivity, implicating aberrant microstructural organisation in the pathophysiology of the disinhibition.^47^ This suggests that elevated neurite density in limbic and associative cortical territories may confer heightened susceptibility to stimulation-induced perturbations within circuits mediating conflict-based decision-making processes. The STN serves as a critical node in the hyperdirect pathway, mediating the rapid suppression of prepotent responses during situations of response conflict.^48^ Individuals with denser neurite architecture in connected cortical territories may exhibit altered propagation of stimulation effects throughout cortico-basal ganglia-thalamo-cortical loops, potentially amplifying the disinhibitory consequences of STN modulation. This interpretation aligns with computational models suggesting that microstructural properties influence the spatiotemporal dynamics of neural signal transmission and, consequently, the efficacy of inhibitory control mechanisms.

Notably, this pattern contrasts with the associations observed in WM, where higher ODI – reflecting disorganised or dispersed fibre architecture – was linked to worse outcomes, whereas intact, coherent fibre bundles appeared to confer protection against treatment-emergent impulsive behaviours. These differential vulnerability patterns likely reflect the fundamentally distinct microstructural properties of grey versus WM: while efficient signal transmission in WM depends on fibre organisation and coherence, grey matter function is governed by the complexity of neuronal circuitry and the balance of excitatory and inhibitory networks.^49^ Higher cortical NDI in our sample may therefore not simply represent “healthy” tissue, but rather hyperreactive areas that become dysregulated following STN modulation.^49 50^ While these findings implicate elevated NDI as a potential vulnerability marker, it is important to recognise that NDI remains a proxy measure of neurite density that does not capture the functional capacity or qualitative characteristics of the underlying neural tissue.^49^

### Mechanistic implications for STN-DBS effects

The differential patterns observed between FA, ODI, and NDI metrics provide complementary insights into the microstructural basis of ICB vulnerability.^47^ While FA primarily reflects overall tract integrity and coherence, NODDI parameters offer more specific information about neurite density and orientation dispersion.^47^ ^51^ The predominance of negative associations in ODI in WM and NDI in GM analyses suggests that reduced axonal density in WM and preserved/heightened dendritic complexity in GM in key limbic and executive regions may create a substrate for dysregulated impulse control.^52^ STN-DBS is known to modulate distributed networks beyond the immediate stimulation site, including frontolimbic circuits involved in reward processing and cognitive control.^10^ In patients with preexisting compromised microstructure, our findings suggest that the therapeutic modulation of these networks by STN-DBS may be less effective, or potentially even counterproductive.

### Clinical implications and preoperative risk assessment

Across all metrics examined, significant associations with postoperative changes in ICB were observed. In line with the overall pattern, preoperative microstructural integrity in WM was consistently linked to greater symptom reduction, suggesting that diffusion-based metrics can capture functionally relevant properties of DBS-responsive circuits.

Taken together, these findings have important implications for clinical practice and preoperative counselling. The identification of microstructural markers associated with ICB risk could enable personalised risk stratification, allowing clinicians to provide more informed preoperative counselling and implement targeted monitoring strategies. Patients with microstructural patterns suggestive of increased ICB risk might benefit from more conservative medication reduction protocols, closer behavioural monitoring, and early implementation of behavioural interventions. Furthermore, the anatomical specificity of our findings suggests potential targets for neuromodulation strategies aimed at reducing ICB risk.

## Limitations

First, while NODDI has been validated in various neurological conditions, specific validation in the context of ICBs in PD is limited. Second, our DTI acquisition parameters were constrained to a spatial resolution of 2.0×2.0×2.0 mm³ with 40 diffusion gradients. These specifications, while consistent with established NODDI protocols in previous studies,^12^ represent a balance between acquisition time, spatial resolution, and signal-to-noise ratio. Extended scanning durations pose particular challenges in PD populations due to increased susceptibility to motion artifacts, necessitating this methodological compromise. Third, inherent limitations of open-label trial designs are applicable to this investigation. Although open-label protocols offer certain methodological advantages, they introduce potential confounding factors, including placebo responses that may influence both subjective outcome measures and overall patient well-being. Additionally, patient-reported metrics are subject to reporting bias and may reflect inaccurate symptom recognition or documentation by participants. Furthermore, the heterogeneous nature of ICBs, encompassing various behavioural domains from gambling to hypersexuality, may require more nuanced analytical approaches that consider specific behavioural subtypes. Future studies incorporating longitudinal designs and larger sample sizes will be essential to validate these findings and explore their clinical utility.

## Conclusion

This study demonstrates that microstructural alterations in frontolimbic and executive control networks are associated with the development of ICB following STN-DBS in PD. The convergent evidence from DTI analyses highlights the importance of insular cortex, cingulate circuits, and major association tracts in determining ICB susceptibility. These findings represent an important step toward personalised risk assessment in STN-DBS candidates and may inform the development of targeted interventions to prevent treatment-emergent behavioural complications. Future research should focus on validating these microstructural biomarkers in larger cohorts and exploring their integration into clinical decision-making algorithms.

## Acknowledgement

The authors would like to thank the participants for their active engagement in this study and Stefanie Spriewald for her support in the coordination of the study.

## Data and Code Availability

The data that support the findings of this study are available on request from the corresponding author (PAL). The data is not publicly available due to privacy or ethical restrictions. All tools used for the analysis of MRI data are based on FreeSurfer Version 7.1 (http://surfer.nmr.mgh.harvard.edu/) and FSL 6.0.5.2 (http://www.fmrib.ox.ac.uk/fsl) packages, which are freely available. Scripts for automation were written in tcshell and parts of the statistics were written in Python using the packages numpy, pandas, seaborn, matplotlib, nibabel and scipy, which are also freely available. Python program code for the analysis of NODDI-DTI is available from https://github.com/dicemt/DTI-NODDI.

## Contributorship

PAL: study concept and design, data acquisition, data analysis, drafting of the manuscript

MN: data acquisition, drafting of the manuscript

LW: data analysis, drafting of the manuscript

LC: data analysis, drafting of the manuscript

AC: data acquisition, critical revision of the manuscript

MBo: data acquisition, surgical intervention, critical revision of the manuscript

AR: critical revision of the manuscript

MH: data acquisition, critical revision of the manuscript

JW: data acquisition, critical revision of the manuscript

CN: data acquisition, surgical intervention, critical revision of the manuscript

KRC: critical revision of the manuscript

HSD: study design, critical revision of the manuscript

LT: study design, critical revision of the manuscript

DJP: data acquisition, drafting of the manuscript

MBe: study concept and design, data acquisition, data analysis, drafting of the manuscript

