## supplementary material for "Microstructure predicts impulsive and compulsive behaviour following subthalamic stimulation in Parkinson’s disease"

**Supplementary Methods**

**MRI Data Acquisition and Processing**

PD patients in the ON-medication state were scanned at baseline on a 3T Trio scanner (Siemens, Erlangen, Germany) at the Core Unit Brain Imaging of the University of Marburg. The acquisition protocol comprised the following sequences:

1. 3D T1-weighted Magnetization Prepared - Rapid Gradient Echo sequence (MPRAGE, field of view (FoV) = 256 mm, matrix 256 x 256, 176 slices, slice thickness 1 mm, voxel dimension 1.0 x 1.0 x 1.0 mm³, repetition time (TR) = 1900 ms, echo time (TE) = 2.26 ms, inversion time (TI) = 900 ms, flip-angle = 9°, bandwidth (BW) = 200 Hz/Pixel, parallel imaging (GRAPPA) with factor 2)
2. Diffusion weighted imaging (DWI) (FoV = 256 mm, matrix 128 x 128, slice thickness 2 mm, distance factor 0 %, voxel dimension 2.0 x 2.0 x 2.0 mm³, TR = 7900 ms, TE = 90 ms, BW = 1502 Hz/Pixel, 40 diffusion encoding gradients, three intermittent non-weighted b0 images (b = 0 s/mm²), high b-value b = 1000 s/mm², GRAPPA with factor 2)

All images were investigated to be free of motion or ghosting and high frequency and/or wrap-around artefacts at the time of image acquisition.

**Supplementary Results**

**Supplementary Table 1**

**Association between FA-values and postoperative Impulsive-Compulsive Outcomes**

| **Fractional Anisotropy** | | |  |  |  |  | | |
| --- | --- | --- | --- | --- | --- | --- | --- | --- |
| Negative  Cluster | Location | Slope | Intercept | p-value | Volume in mm³ | MNI152-Coordinates | | |
|  |  |  |  |  |  | X | Y | Z |
| N1 | Left angular Gyrus | -.0005 | .1506 | < .001 | 419 | -47 | -51 | 15 |
| N2 | Right paracingulate gyrus | -.0003 | .1378 | < .001 | 293 | 4 | 37 | -9 |
| N3 | Left V | -.0002 | .2024 | < .001 | 260 | -16 | -40 | -20 |
| N4 | Left insular cortex  Uncinate fasciculus  Inferior fronto- occipital fasciculus | -.0003 | .4115 | < .001 | 232 | -22 | 8 | -11 |
| N5 | Right putamen  Uncinate fasciculus  Inferior fronto- occipital fasciculus | -.0002 | .1992 | < .001 | 215 | 25 | 5 | -5 |
| N6 | Left lingual gyrus | -.0003 | .1725 | < .001 | 213 | -22 | -51 | -4 |
| N7 | Right crus I  Right VI | -.0002 | .1677 | < .001 | 188 | 22 | -73 | -18 |
| Positive Cluster |  |  |  |  |  |  |  |  |
| P1 | Left cingulum | .0003 | .4918 | < .001 | 306 | -20 | -56 | 39 |

**Supplementary Table 1.** Characteristics of clusters with an association between PD patients’ FA-values and postoperative change in QUIP-RS. “Positive Cluster” denotes clusters with a positive association between patients’ FA-values and percentage differences in QUIP-RS, i.e. higher FA-values were associated with higher postoperative change. “Negative Cluster” denotes clusters with a negative association between patients’ FA-values and percentage difference of QUIP-RS, i.e. higher FA-values were associated with lower postoperative change. “Location” indicates the anatomical landmark comprising the majority of voxels of a cluster according to Johns Hopkins University (JHU) white matter atlas, Harvard-Oxford cortical and subcortical atlas, and University College London (UCL) cerebellar atlas. P-Values are clusterwise p-values corrected for multiple comparisons. “Volume in mm³” denotes the size of a cluster and “MNI152-coordinates” describes the coordinates of the cluster’s center of gravity in MNI152-space.

**Supplementary Table 2**

**Association between ODI-values and postoperative Impulsive-Compulsive Outcomes**

| **Orientation Dispersion Index** | | |  |  |  |  | | |
| --- | --- | --- | --- | --- | --- | --- | --- | --- |
| Negative  Cluster | Location | Slope | Intercept | p-value | Volume in mm³ | MNI152-Coordinates | | |
|  |  |  |  |  |  | X | Y | Z |
| N1 | Left WM adjacent to superior parietal lobule  Left cingulum | -.0002 | .1267 | < .001 | 421 | -21 | -47 | 40 |
| N2 | Right WM adjacent to frontal pole  Forceps minor | -.0002 | .2064 | < .001 | 267 | 21 | 43 | 11 |
| N3 | Right WM adjacent to frontal pole  Superior frontal gyrus  Forceps minor | -.0002 | .1947 | < .001 | 229 | 19 | 34 | 27 |
| N4 | Left WM adjacent to superior frontal gyrus | -.0002 | .2024 | < .001 | 210 | -20 | 12 | 37 |
| N5 | Left WM adjacent to postcentral gyrus  Corticospinal tract  Superior longitudinal fasciculus | -.0002 | .1550 | < .001 | 175 | -30 | -32 | 40 |
| N6 | Right inferior fronto-occipital fasciculus | -.0002 | .2016 | .001 | 171 | 27 | 29 | 3 |
| N7 | Left cingulum | -.0004 | .3388 | < .001 | 162 | -12 | 26 | 26 |
| N8 | Left WM adjacent to frontal pole and superior frontal gyrus  Forceps minor | -.0002 | .1939 | < .001 | 157 | -16 | 35 | 25 |
| N9 | Left WM adjacent to insular cortex  Superior longitudinal fasciculus  Uncinate fasciculus | -.0004 | .2221 | < .001 | 157 | -34 | 2 | 1 |
| N10 | Left WM adjacent to supramarginal gyrus  Superior longitudinal fasciculus | -.0002 | .1965 | < .001 | 155 | -44 | -44 | 14 |
| N11 | Left WM adjacent to frontal orbital cortex and frontal pole  Uncinate fasciculus | -.0003 | .2579 | < .001 | 150 | -21 | 31 | -9 |
| N12 | Right WM adjacent to middle temporal gyrus  Inferior longitudinal fasciculus | -.0003 | .2492 | < .001 | 147 | 47 | -12 | -19 |
| N13 | Right inferior temporal gyrus  Middle temporal gyrus  Superior longitudinal fasciculus | -.0005 | .2861 | < .001 | 143 | 52 | -54 | -7 |
| N14 | Left middle temporal gyrus  Superior longitudinal fasciculus | -.0003 | .2026 | < .001 | 140 | 49 | -38 | -2 |
| N15 | Left WM adjacent to precuneous cortex  Cingulum | -.0003 | .2336 | < .001 | 138 | -13 | -52 | 15 |
| N16 | Left WM adjacent to superior temporal gyrus  Inferior longitudinal fasciculus | -.0003 | .1762 | < .001 | 138 | -46 | -22 | -5 |
| N17 | Left WM adjacent to inferior frontal gyrus | -.0002 | .2270 | < .001 | 127 | -32 | 14 | 21 |
| N18 | Left WM adjacent to precentral gyrus  Superior longitudinal fasciculus | -.0002 | .1819 | < .001 | 127 | -40 | -3 | 21 |
| Positive Cluster |  |  |  |  |  |  |  |  |
| P1 | Left temporal fusiform cortex | .0006 | .3197 | < .001 | 162 | -45 | -41 | -17 |

**Supplementary Table 2.** Characteristics of clusters with an association between PD patients’ ODI-values and postoperative change in QUIP-RS. “Positive Cluster” denotes clusters with a positive association between patients’ ODI-values and percentage differences in QUIP-RS, i.e. higher ODI-values were associated with higher postoperative change. “Negative Cluster” denotes clusters with a negative association between patients’ ODI-values and percentage difference of QUIP-RS, i.e. higher ODI-values were associated with lower postoperative change. “Location” indicates the anatomical landmark comprising the majority of voxels of a cluster according to Johns Hopkins University (JHU) white matter atlas, Harvard-Oxford cortical and subcortical atlas, and University College London (UCL) cerebellar atlas. P-Values are clusterwise p-values corrected for multiple comparisons. “Volume in mm³” denotes the size of a cluster and “MNI152-coordinates” describes the coordinates of the cluster’s center of gravity in MNI152-space.

**Supplementary Table 3**

**Association between NDI-values and postoperative Impulsive-Compulsive Outcomes**

| **Neurite Density Index** | | |  |  |  |  | | |
| --- | --- | --- | --- | --- | --- | --- | --- | --- |
| Negative  Cluster | Location | Slope | Intercept | p-value | Volume in mm³ | MNI152-Coordinates | | |
|  |  |  |  |  |  | X | Y | Z |
| N1 | Left middle temporal gyrus  Inferior longitudinal fasciculus | -.0004 | .4953 | < .001 | 346 | -45 | -22 | -6 |
| N2 | Left inferior frontal gyrus | -.0005 | .5374 | < .001 | 270 | -32 | 20 | 16 |
| N3 | Right paracingulate gyrus  Forceps minor | -.0005 | .4255 | < .001 | 251 | 14 | 36 | -6 |
| N4 | Left Insular cortex  Uncinate fasciculus | -.0004 | .3501 | < .001 | 223 | -34 | 9 | -6 |
| N5 | Left precentral gyrus  Superior longitudinal fasciculus | -.0004 | .5579 | < .001 | 171 | -37 | 1 | 21 |
| N6 | Left temporal pole  Inferior longitudinal fasciculus  Uncinate fasciculus | -.0004 | .4442 | < .001 | 170 | -42 | 8 | -25 |
| N7 | Left paracingulate gyrus | -.0006 | .5748 | < .001 | 154 | -14 | 26 | 26 |
| N8 | Right inferior frontal gyrus (pars opercularis) and adjacent WM | -.0004 | .5898 | < .001 | 144 | 34 | 17 | 14 |
| N9 | Left supplementary motor cortex | -.0004 | .5799 | < .001 | 141 | -15 | -6 | 42 |
| N10 | Right I-IV | -.0007 | -.3428 | < .001 | 135 | 1 | -55 | -8 |
| N11 | Right brainstem nuclei, mesencephalon | -.0004 | .6278 | < .001 | 130 | 7 | -38 | -21 |
| N12 | Right intracalcarine cortex | -.0008 | .4191 | < .001 | 128 | 14 | -71 | 17 |
| N13 | Left putamen | -.0006 | .4395 | < .001 | 124 | -29 | 0 | -7 |

**Supplementary Table 3.** Characteristics of clusters with an association between PD patients’ NDI-values and postoperative change in QUIP-RS. “Positive Cluster” denotes clusters with a positive association between patients’ NDI-values and percentage differences in QUIP-RS, i.e. higher NDI-values were associated with higher postoperative change. “Negative Cluster” denotes clusters with a negative association between patients’ NDI-values and percentage difference of QUIP-RS, i.e. higher NDI-values were associated with lower postoperative change. “Location” indicates the anatomical landmark comprising the majority of voxels of a cluster according to Johns Hopkins University (JHU) white matter atlas, Harvard-Oxford cortical and subcortical atlas, and University College London (UCL) cerebellar atlas. P-Values are clusterwise p-values corrected for multiple comparisons. “Volume in mm³” denotes the size of a cluster and “MNI152-coordinates” describes the coordinates of the cluster’s center of gravity in MNI152-space.

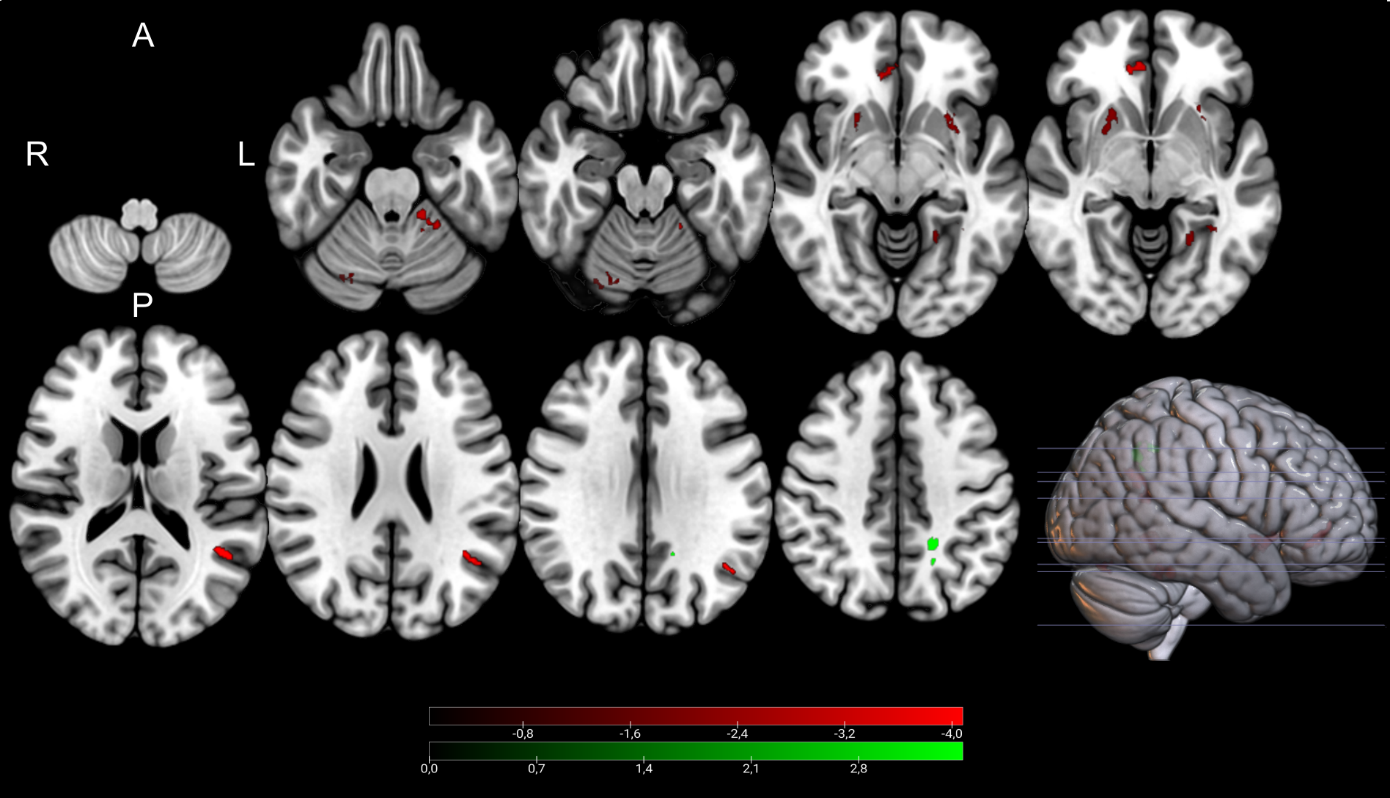

**Supplementary Figure 1.** Clusters with a positive (green) and negative (red) association between PD patients’ FA-values and postoperative change in QUIP-RS, as revealed by the whole brain analysis. P-Values were corrected for multiple comparisons using a permutation-based approach and shown as the negative decadic logarithm of the p-value.
